# Use of repeated mammograms to evaluate risk of breast cancer: a systematic review of methods used in the literature

**DOI:** 10.1101/2021.11.10.21266200

**Authors:** Akila Anandarajah, Yongzhen Chen, Carolyn Stoll, Angela Hardi, Shu Jiang, Graham A. Colditz

## Abstract

**Objective:** This systematic review aimed to assess methods used to relate repeated mammographic images to breast cancer risk, including the time from mammogram to diagnosis of breast cancer, and methods for analysis of data from either one or both breasts (averaged or assessed individually).

**Design:** A systematic review was performed.

**Setting:** The databases including Medline (Ovid) 1946-, Embase.com 1947-, CINAHL Plus 1937-, Scopus 1823-, Cochrane Library (including CENTRAL), and Clinicaltrials.gov were searched through October 2021 to extract published articles in English describing the relationship of change in mammographic features with risk of breast cancer.

**Participants:** Women with mammogram images.

**Main outcome measure:** Breast cancer incidence.

**Results:** Twenty articles were included in the final review. We found that BIRADs and Cumulus were most commonly used for classifying mammographic density and automated assessment was used on more recent digital mammograms. Time between mammograms varied from 1 to median of 4.1 years, and only 9 of the studies used more than 2 mammograms to quantify features. One study used a prediction horizon of 5 and 10 years, one used 5 years only and another 10 years only, while in the others the prediction horizon was not clearly defined with investigators using the next screening mammogram.

**Conclusion:** This review provided an updated overview of the state of the art and revealed research gaps; based on these, we provide recommendations for future studies using repeated measure methods for mammogram images to make the use of accumulating image data. By following these recommendations, we expect to improve risk classification and risk prediction for women to tailor screening and prevention strategies to level of risk.

**Article summary:** *Strengths and limitations of the study:*

- To the best of our knowledge, this is the most recent systematic review on the topic of using multiple mammogram images to define risk of breast cancer.
- This review was performed strictly following systematic review guidelines including a medical librarian with expertise in searching, multiple independent reviewers involved in study selection and data extraction, and reporting following PRISMA 2020 guidelines.
- Due to heterogeneity of methods for assessment and classification (categorical and continuous) of mammographic features including breast density and time to breast cancer, we did not perform risk of bias or conduct a meta-analysis.
- Few studies looked at repeated measures of non-density features.

## Introduction

Evolving technology from film mammograms to digital images has changed the sources of data and ease of access to study a range of summary measures from breast mammograms and risk of breast cancer.^1^ In particular, given women have repeated mammograms as part of a regular screening program,^2-4^ access to repeated images has become more feasible in real time for risk classification. Improved risk classification is fundamental to counseling women for their risk management.^5 6^

The leading measure for risk categorization extracted from mammograms is breast density.^7 8^ This is now widely used and reported with many states requiring return of mammographic breast density measures to women as part of routine screening. Mammographic breast density is a strong reproducible risk factor for breast cancer across different approaches used to measure it (clinical judgement or semi/automated estimation).^7^ Mammographic breast density has typically been measured as an average value across both left and right breasts to relate to risk of subsequent breast cancer. Change in breast density has been much less frequently studied. However, growing access to the large data from mammograms encourages a reassessment of the approaches employed to assess change in density and risk of subsequent breast cancer.^9 10^

Guidelines recommend screening mammography from age 45 (American Cancer Society^2^) or 50 (US Preventive Services Task Force^3^), with either annual or biennial mammography.^4^ Women generally have a series of repeated mammograms (longitudinal data). Additionally, these recurring screening mammograms capture both the left and right breast (bivariate profiles). See Figure 1. Despite the availability of bivariate longitudinal images, general decision making is still based on mammographic breast density at a point in time, averaged between the two breasts,^11^ to forecast the overall breast cancer risk. While a growing number of studies use more than just baseline mammogram values which could improve risk classification and is promising for clinical decision making, we note there is no systematic review and summary of these studies, although a recent publication reported results from 9 studies and combined results showing a positive association between increase in BIRADs density category and increase in breast cancer risk.^12^ A richer summary of methods used to classify density and other features on mammograms and evaluate change in relation to risk can identify common approaches and help guide the use of change for breast cancer risk prediction. Therefore, we undertook the current systematic review.

**Figure 1.**
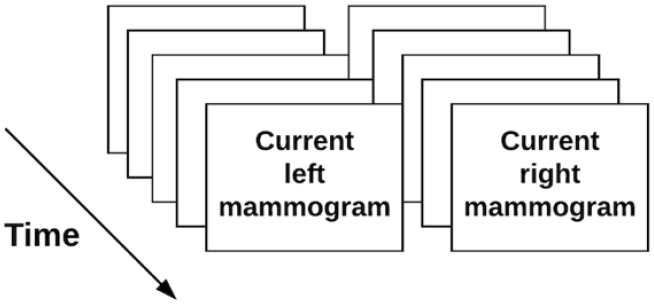
Illustration of the available mammography data from routine screening.

We aim to summarize the methods used, the time from mammogram to diagnosis of breast cancer, methods for analysis of data from either one or both breasts (averaged or assessed individually), and identify gaps in evidence to prioritize future studies.

## Materials and Methods

### Eligibility Criteria

#### Population

We considered all studies of adult women (at least eighteen years old) involving original data. Abstract-only papers, review articles, and conference papers were excluded. Intervention: We included studies measuring change in mammographic features between mammograms. A study had to use at least two different mammograms to be included.

#### Outcomes

Our primary outcomes of interest were risk of breast cancer, including both invasive and in situ cancers, and differences in mammographic features over time. Risk of breast cancer was required to be dichotomized (yes/no), and analysis of other risks (e.g., risk of interval vs. screen-detected cancer) were not included. Studies were required to assess the relationship of the change in mammographic features with risk of breast cancer.

Only studies available in English were included.

### Information Sources

The published literature was searched using strategies designed by a medical librarian (AH) for the concepts of breast density, mammography, and related synonyms. These strategies were created using a combination of controlled vocabulary terms and keywords, and were executed in Medline (Ovid) 1946-, Embase.com 1947-, CINAHL Plus 1937-, Scopus 1823-, Cochrane Library (including CENTRAL), and Clinicaltrials.gov. Results were limited to English using database-supplied filters. Letters, comments, notes, and editorials were also excluded from the results using publication type filters and limits.

### Search Strategy

An example search is provided below (for Embase).

(‘breast density’/exp OR ((breast NEAR/3 densit*):ti,ab,kw OR (mammary NEAR/3 densit*):ti,ab,kw OR (mammographic NEAR/3 densit*):ti,ab,kw)) AND (‘mammography’/deOR mammograph*:ti,ab,kwOR mammogram*:ti,ab,kwOR mastrography:ti,ab,kwOR ‘digital breast tomosynthesis’:ti,ab,kwOR ‘x-ray breast tomosynthesis’:ti,ab,kw)NOT(‘editorial’/it OR ‘letter’/it OR ‘note’/it) AND [english]/lim

The search was completed for the first time on September 9, 2020, and was run again on October 14, 2021 to retrieve citations that were published since the original search. The second search was dated limited to 2020-present. Full search strategies are provided in the appendix.

### Selection Process

Two reviewers (AA, CS) worked independently to review the titles and abstracts of the records. Next, the two reviewers independently screened the full text of the articles that they did not reject and indicated those measuring mammographic features over time, which were ultimately eligible for inclusion. Any disagreements of which articles to include were resolved by consensus.

Reference lists of included studies were hand searched to find additional relevant studies.

### Data Collection Process

We created a data extraction sheet which two reviewers (AA, YC) used to independently extract data from the included studies. Disagreements were resolved by a third reviewer. If included studies were missing any desired information, any additional papers from the work cited, such as previous reports, methods papers, or protocols, were reviewed for this information.

### Data Items

Any estimate of change in a mammographic feature over time or risk of breast cancer was eligible to be included. Predictive ability could be evaluated using an area under the curve, hazard ratio, odds ratio, relative risk, 5-year risk, or p value. Change could be reported as a percentage or an absolute value. No restrictions on follow-up time were placed. For studies that reported multiple risk estimates, we prioritized the primary models which were discussed in the results section of the paper. If all models were discussed equally, then we listed the models with the best ability to predict breast cancer. For studies that reported multiple types of change, we prioritized the primary types which were discussed in the results section of the paper. If all types were discussed equally, then we listed the most frequent types of change.

We collected data on:

the report: author, publication year

the study: location/institution, number of cases, number of controls

the research design and features: lapsed time from mammogram to diagnosis

the mammogram: machine type, mammogram view(s), breast(s) used for analysis, time between mammograms, number of mammograms

the model: how density was measured, type of model, baseline variable(s), texture feature(s), prediction horizon

### Risk of Bias

The objective of this review is to summarize the methods and analysis techniques used to assess change in mammographic features and risk of breast cancer rather than to quantitatively synthesize the results of the studies. Therefore, a risk of bias assessment, while typically performed in a systematic review, would not serve the objective and was not performed.

### Patient and public involvement

No patient involvement

### Human subjects

This study did not involve human subjects and therefore oversight from an Institutional Review Board was not required.

### Registration and protocol

This review was not registered and a protocol was not prepared.

## Results

The search and study selection process is shown in Figure 2. A total of 11,111 results were retrieved from the initial database literature search and imported into Endnote. 11 citations from ClinicalTrials.gov were retrieved and added to an Excel file library. After removing duplicates 4,863 unique citations remained for analysis. The search was run again in October 2021 to retrieve citations that were published since the original search. A total of 1,633 results were retrieved and imported to Endnote. After removing duplicates, including duplicates from the original search, 466 unique citations were added to the pool of results for analysis. Between the two searches a total of 11,577 results were retrieved, and there were 5,329 unique citations.

**Figure 2:**
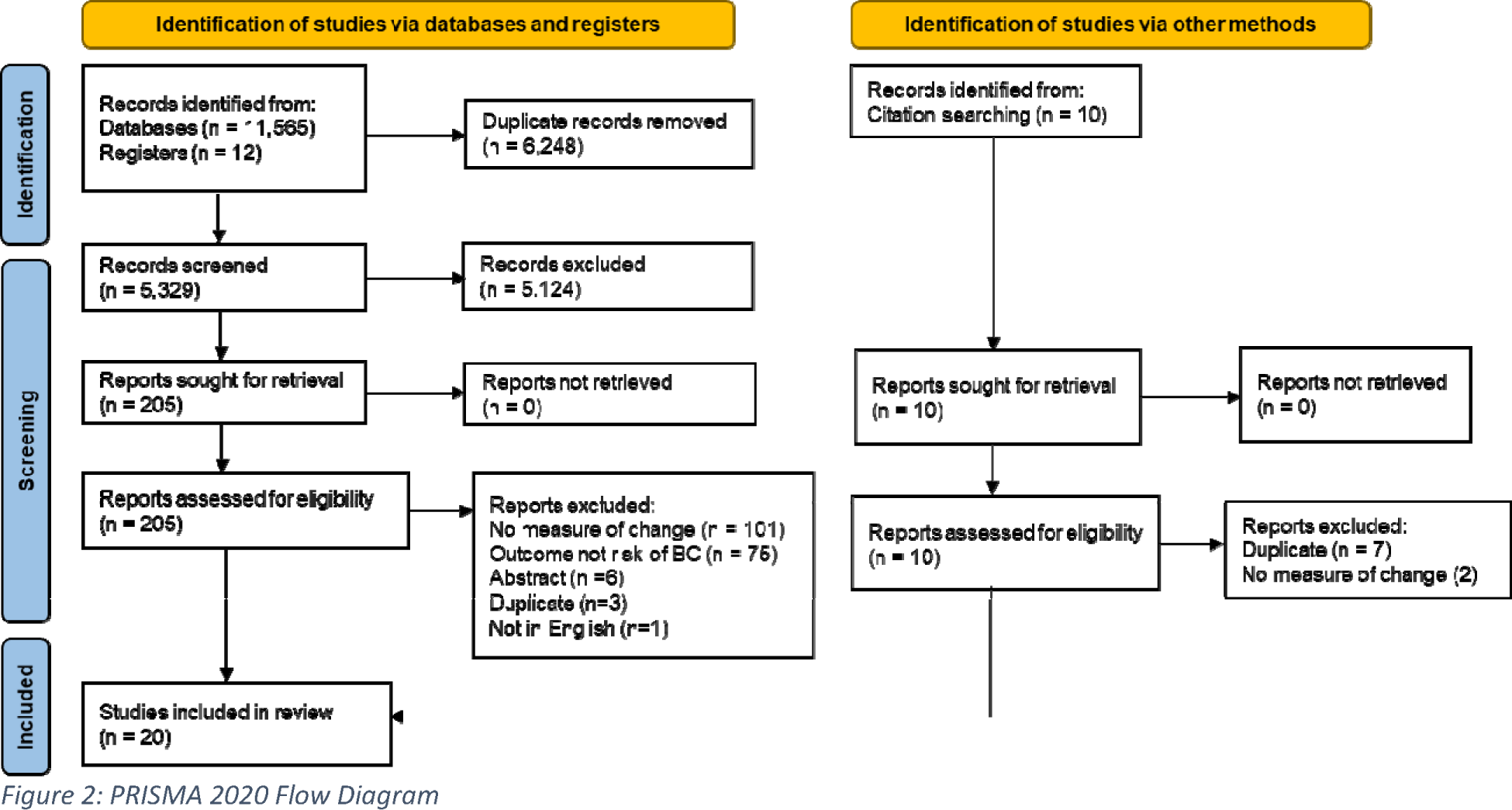
PRISMA 2020 Flow Diagram

Of the 5,329 unique citations, 5,124 were excluded based on review of title and abstract. 205 full-text reports were retrieved and assessed for eligibility by two readers. Of these, 186 were excluded for reasons such as not measuring change, not having risk of breast cancer as an outcome, being an abstract or a duplicate paper, or not being published in English.

10 potential reports were identified from hand searching of citations. All of these were reviewed by full-text, and 9 were excluded for being duplicates or not having a measure of change in a mammographic feature.

After fully screening search results, 20 studies meeting eligibility criteria were included in the review.^13-32^ These 20 studies used 2 or more mammograms to relate change in density or other features to risk of breast cancer and met eligibility criteria as set out in the selection flow chart. See PRISMA flow chart (Figure 2).

The key descriptive features of the 20 eligible studies are summarized in table 1. These rely on mammography film records (10 studies) though studies published from 2016 onwards often use digital images. Number of cases included in each study varied from a low of 45 cases^18^ to a high of 1592 in a Spanish case control study.^23^ See table 1.

**Table 1.** Studies using repeated assessment of mammographic features included in systematic review (sorted by year published)

| Author | Year | Location/<br>institution | Machine type | View used<br>(CC/MLO/both) | # cases | # controls | Time from mmg to diagnosis<br>of cancer |
| --- | --- | --- | --- | --- | --- | --- | --- |
| Salminen<sup>27</sup> | 1998 | The Cancer Society of Finland | film | NR | 68 | 4013 | At least 6 months |
| van Gils<sup>31</sup> | 1999 | Nijmegen, Netherlands | film | Up until 1981/1982 the lateromedial view was used and, after that time, the MLO view | 108 | 400 | Prior to diagnosis. Diagnosed with cancer between 1985 and 1994. Screening from 1975-1994. |
| Maskarinec<sup>25</sup> | 2006 | Hawaii and Los Angeles | film | CC | 607 | 667 | The earliest mammogram was taken $6.3 \pm 4.0$ years before diagnosis |
| Kerlikowske<sup>20</sup> | 2007 | Breast Cancer Surveillance Consortium | NR | NR | 2639 | 299316 | Within 12 months of the last examination |
| Vachon<sup>30</sup> | 2007 | Mayo Clinic | film | CC | 372 | 713 | The time interval between the initial mammogram and the diagnosis of cancer or exam date in controls was $7.0 \pm 1.5$ years on average (range, 2.1-10.4). |
| Lokate<sup>24</sup> | 2013 | Utrecht, Netherlands | over 99% of the mammograms were film | MLO | 533 | 1367 | Mammograms taken until diagnosis |
| Work<sup>32</sup> | 2014 | Columbia University Medical Center | film | NR | 85 | 85 | Cancer diagnosis occurred a median of 1.5 years after the second mammogram (range, 6 months to 9.4 years) |
| Kerlikowske<sup>19</sup> | 2015 | Breast Cancer Surveillance Consortium | NR | NR | 13715 | 708939 | Diagnosed during follow-up period (which has mean = 6.6 years, range = 1 day to 10 years). Women diagnosed the 3 months following their second examination were excluded. |
| Tan<sup>29</sup> | 2016 | University of Pittsburgh Medical Center | digital (not specified further) | both | 159 | 176 | The average elapsed time between the “current” and each of “prior” #1, #2 and #3 studies was 1.16±0.41, 2.30±0.55 and 3.44±0.72 years, respectively. |
| Busana<sup>15</sup> | 2016 | National Health Service | film | MLO | 313 | 452 | Only images taken at least 1 year prior to diagnosis were included |
| Humphrey<sup>17</sup> | 2016 | Manchester, England | digital (not specified further) | both | 170 | 510 | Cases had an initial negative mammographic screen and another three years later when cancer was diagnosed. |
| Khoo<sup>21</sup> | 2016 | Sweden | film | MLO | 250 | 250 | Mammogram before diagnosis used |
| Byrne<sup>16</sup> | 2017 | Women’s Health Initiative | film | CC | 174 | 733 | NR |
| Brandt<sup>14</sup> | 2019 | Mayo Clinic Rochester and the San Francisco Mammography Registry | Hologic Selenia | both | 1160 | 2360 | Within 2 months and 1–5 years before diagnosis |
| Román<sup>26</sup> | 2019 | Spain | film and digital (not specified further) | both | 1592 | 115796 | Diagnosed within two years of the last screening examination in the study period |
| Azam<sup>13</sup> | 2020 | Sweden | GE Medical Systems, Philips Healthcare, Sectra Imtec AB and FUJI | MLO | 563 | 43247 | Diagnosed during follow-up period (which has average = 5.4 years) |
| Kim<sup>22</sup> | 2020 | Kangbuk Samsung Hospital Total Healthcare Centers | Hologic Selenia Dimensions and General Electric Senographe 2000D/DMR/DS A21 | both | 803 | 73446 | Diagnosed during follow-up period (which has median = 6.1 years, interquartile range = 4.1-8.8 years, maximum = 13 years) |
| Sartor<sup>28</sup> | 2020 | Skane University Hospital | Approximately 90% of all images were acquired on Siemens | both | 51 | 102 | The last mammogram was defined as the last screening round before diagnosis (76 %) or the diagnostic mammographic examination |
|  |  |  | mammography systems: 21% on Novation and 69% on Inspiration, with the remaining 10% on GE Senograph |  |  |  | (24%) |
| Kang<sup>18</sup> | 2021 | Samsung Medical Center Health Promotion Center | GE Senograph DS/ESSENTIAL | both | 45 | 3552 | Diagnosed during follow-up period (which has median = 4.8 years, interquartile range 2.8–7.5 years) |
| Kim<sup>23</sup> | 2021 | Korean National Health Insurance Service | NR | NR | 22781 | 3278498 | Excluded women diagnosed with breast cancer within 90□ days after the second screening. Screening in both 2009 to 2010 and 2011 to 2012 with breast cancer incidence identified up to December 2017. |

NR: Not reported

Measures of breast density used in these studies are summarized in Table 2. BIRADS (6 studies) and Cumulus (5 studies) were the most commonly used methods for density assessment. Automated assessments were used on digital mammograms. Table 2 shows that the time between mammograms varied across studies from 1 to median of 4.1 years reflecting differences in guidelines and screening practice across countries. 20 association studies reported time between mammograms of 1 to 3 or more years, and most used only 2 mammogram measures of density or other features. Of note, from the 20 studies only 9 used more than 2 images separated in time to assess change in relation to risk. Furthermore, the covariates used to adjust estimates of association varied substantially across these studies.

**Table 2.**
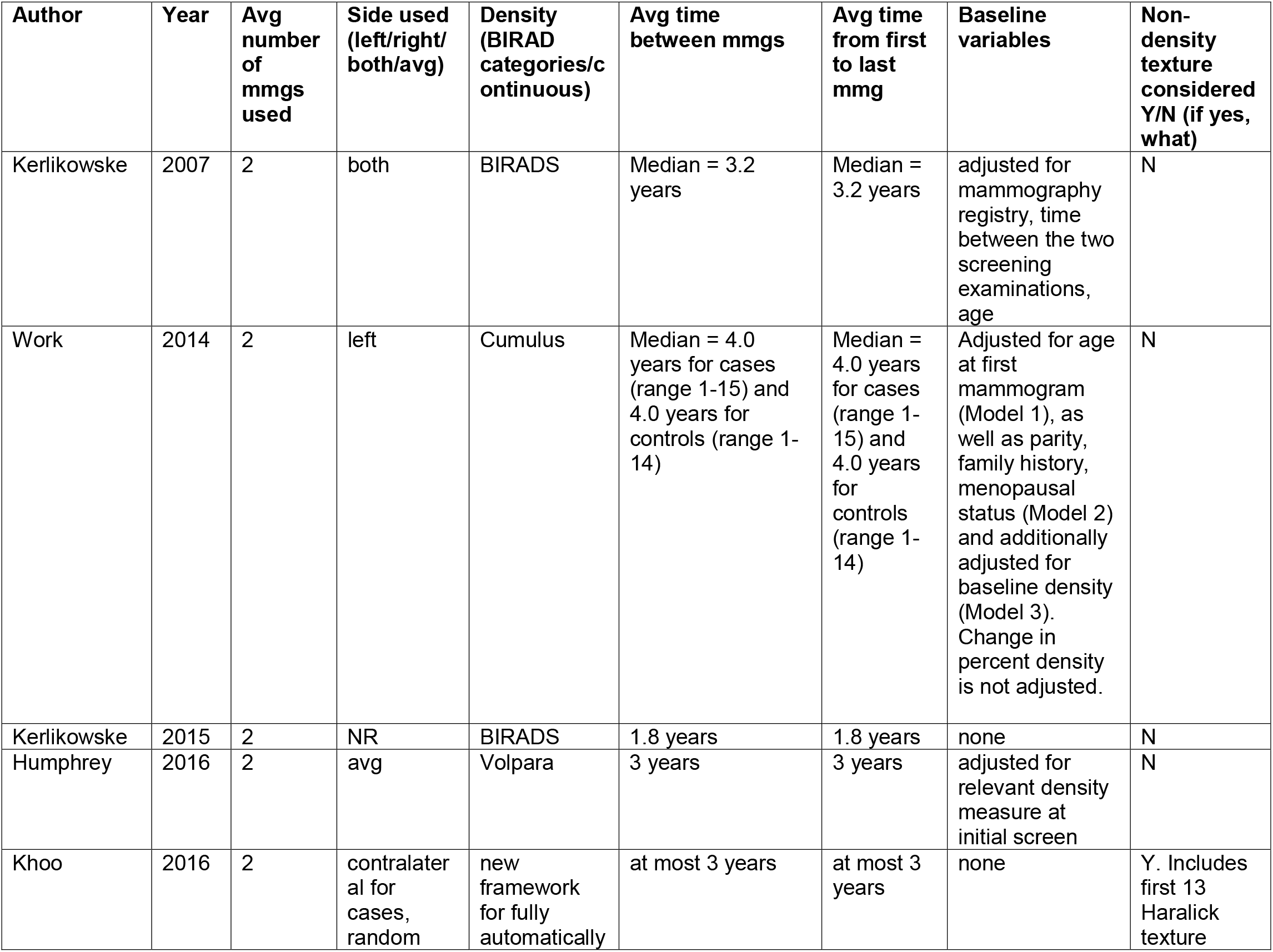
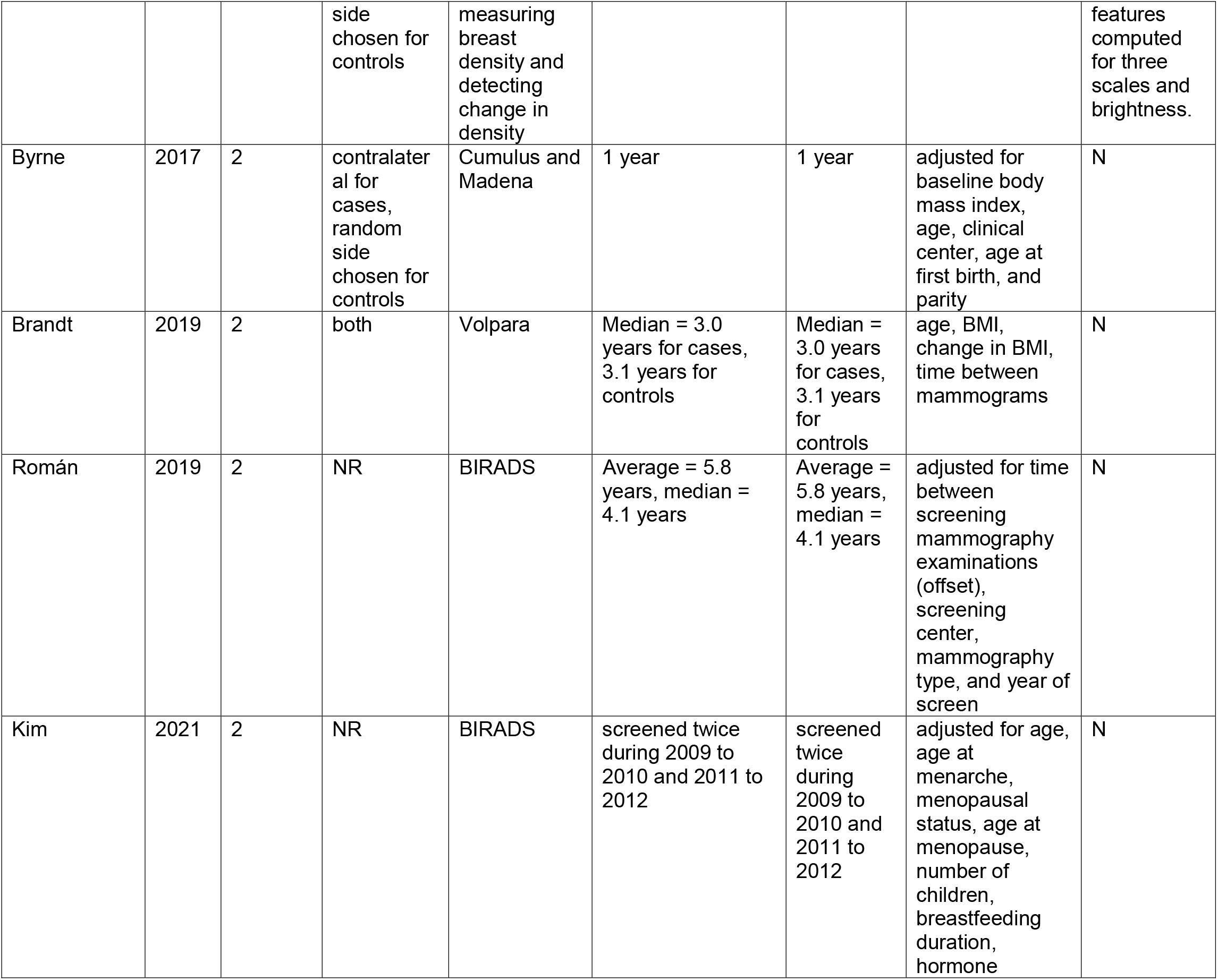
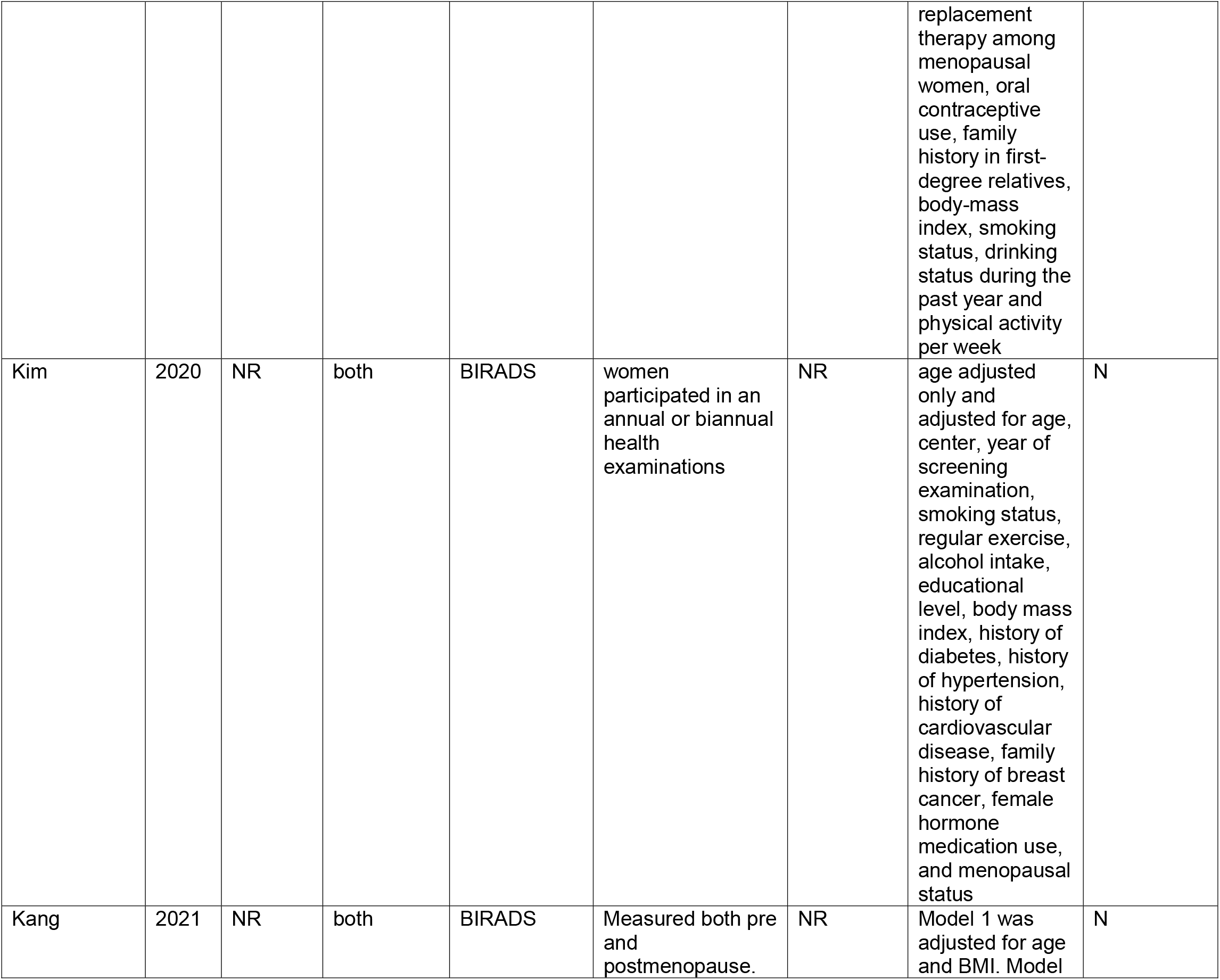
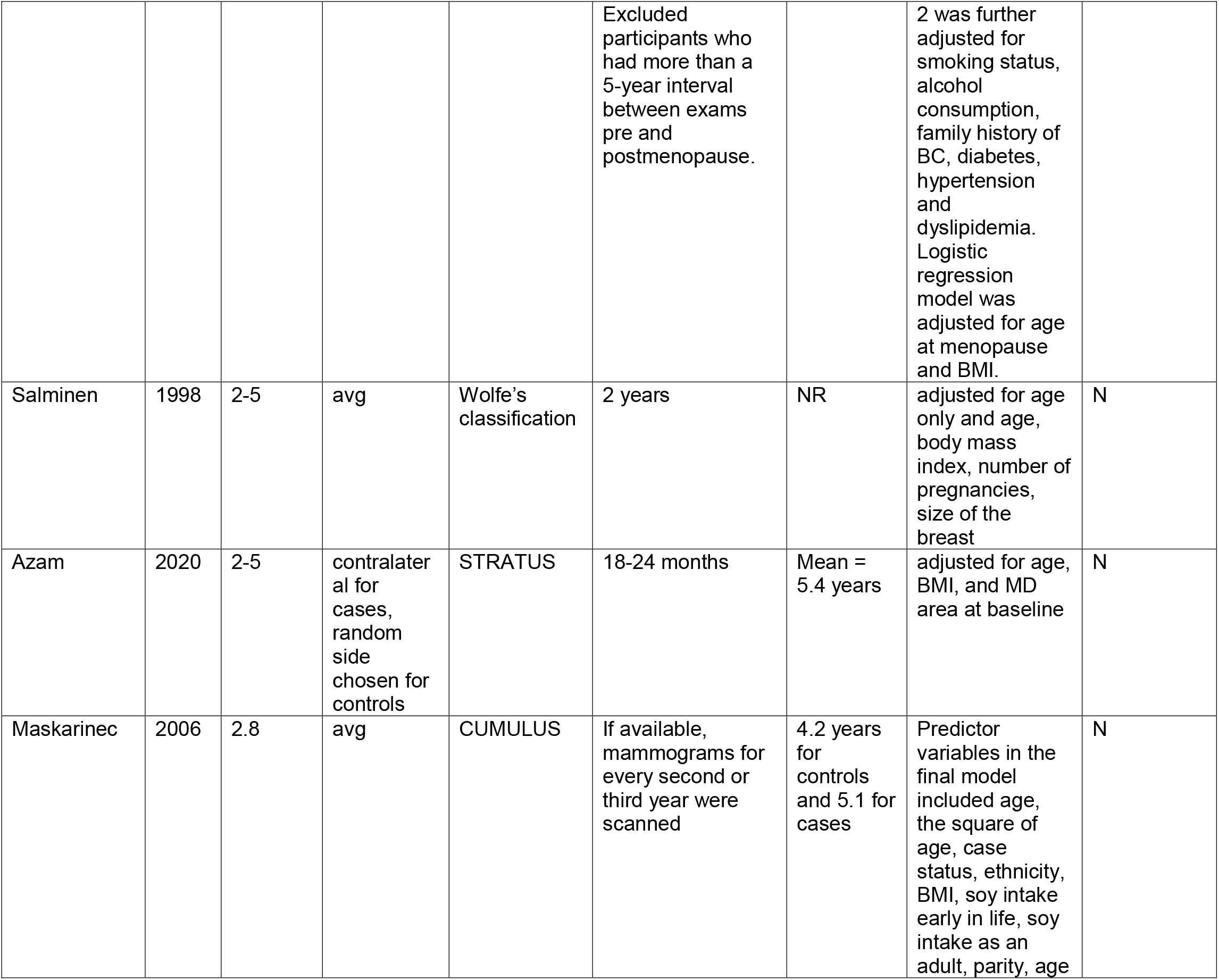
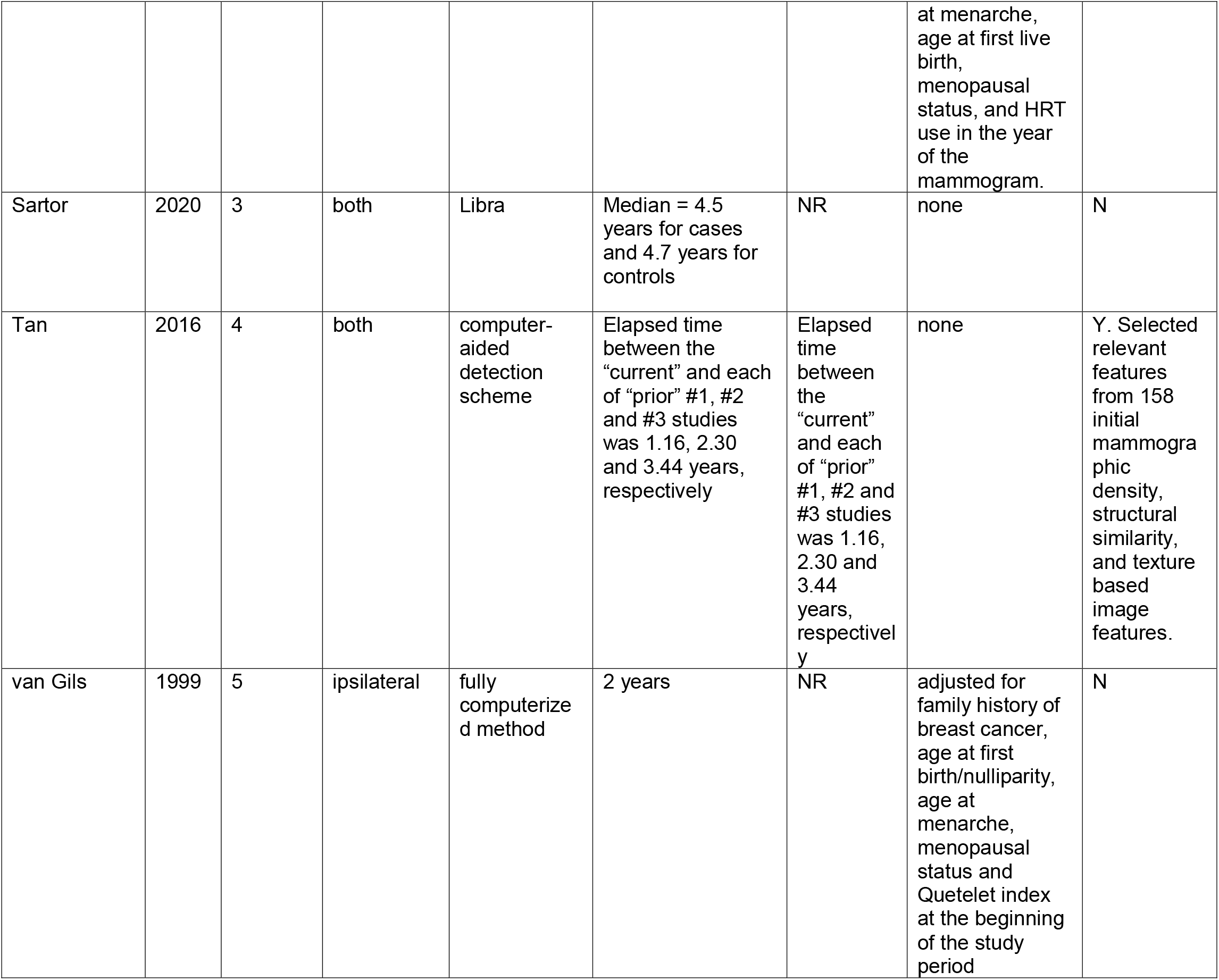
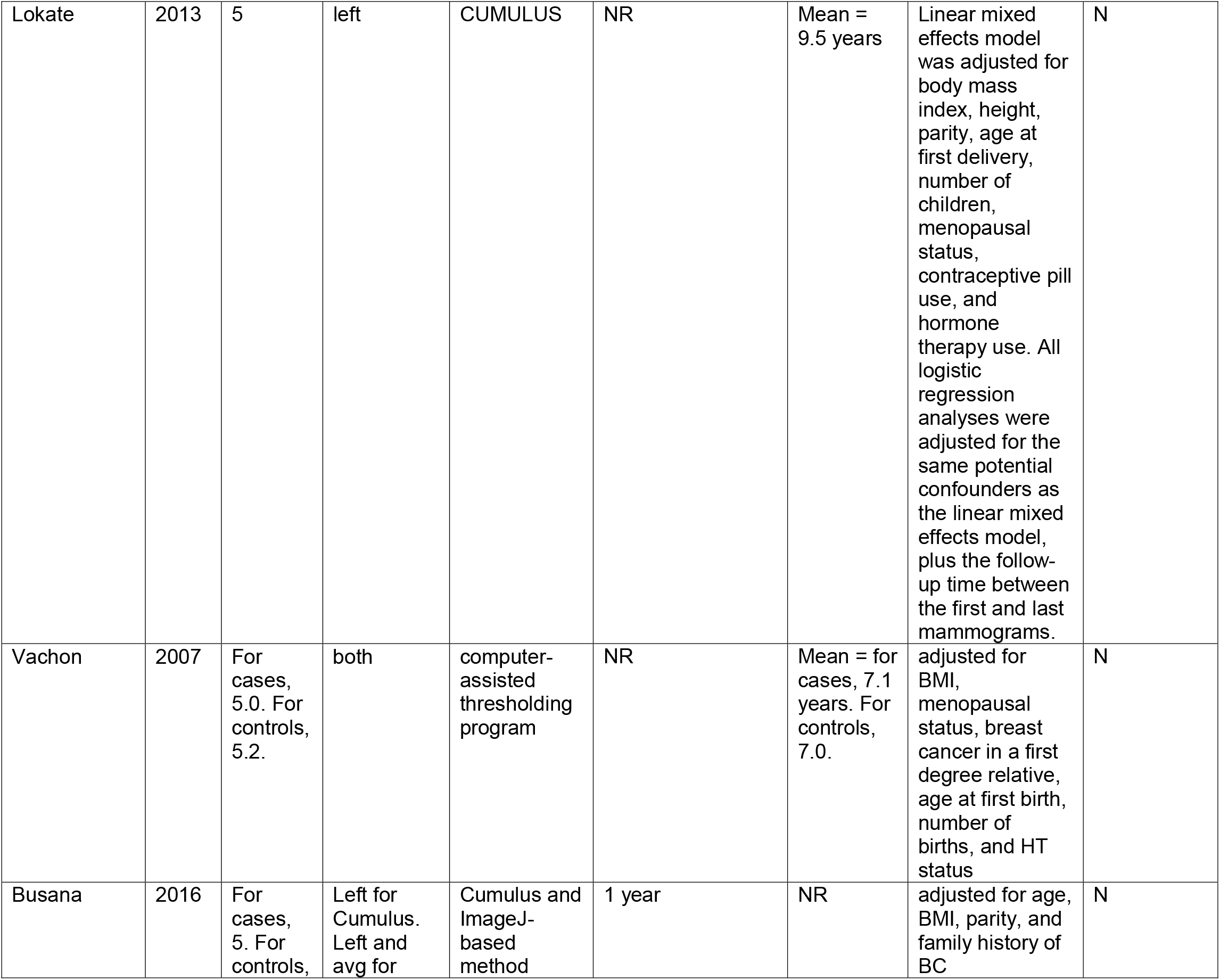
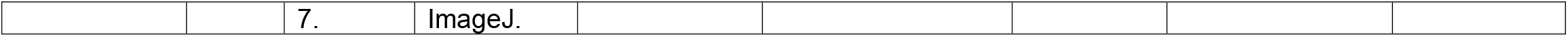
Features of studies using repeated measures of mammographic features and breast cancer risk (sorted by increasing number of mammograms used)

Data from studies of change in mammographic density or features and subsequent risk incorporated into prediction models are summarized in Table 3. Here we also summarize the number of mammograms used and the prediction horizon. Only 1 (Kerlikowske^19^ based on change in BIRADs category between 2 mammograms) reported prediction horizon of 5 and 10 years. In others the prediction horizon was not clearly defined with investigators using the next screening mammogram.^14 21 29 30 33^ There is much variation in approaches to analysis used to relate change in mammographic breast density or features to breast cancer risk. Approaches included change in BIRADs category, change from first to last image (ignoring intermediate images), and change in density as a continuous measure.

**Table 3.** Models of repeated measures of mammographic features and incidence of breast cancer reporting AUC (sorted by number mammograms used)

| Author | Year | Avg number of mmgs used | Model of change (e.g., difference between two densities, mixed effects model, etc.) | Prediction horizon (5/10yr) | AUC (baseline with density) | Overall AUC (with change in density or texture features) | Change in AUC for cases | Change in AUC for controls |
| --- | --- | --- | --- | --- | --- | --- | --- | --- |
| Kerlikowske* | 2015 | 2 | Cox proportional hazards regression and logistic regression | 5 and 10 year | 5-year risk model: 0.635 (0.635 from 5-fold cross-validation). 10-year risk model: 0.622. 5-year risk model among women who changed density categories: 0.630 (0.629 from 5-fold cross-validation). | 5-year risk model: 0.640 (0.639 from 5-fold cross-validation). 10-year risk model: 0.628. 5-year risk model among women who changed density categories: 0.641 (0.639 from 5-fold cross-validation). | A total of 63.5% of women had the same BI-RADS density on two sequential examinations while 17.9% had an increase in breast density category and 18.6% had a decrease. The most common combinations of changing density categories were heterogeneously dense on the earlier examination and scattered fibroglandular on the most recent examination (10.0%) and scattered fibroglandular densities on the earlier examination and heterogeneously dense on the most recent examination (9.9%). | A total of 63.5% of women had the same BI-RADS density on two sequential examinations while 17.9% had an increase in breast density category and 18.6% had a decrease. The most common combinations of changing density categories were heterogeneously dense on the earlier examination and scattered fibroglandular on the most recent examination (10.0%) and scattered fibroglandular densities on the earlier examination and heterogeneously dense on the most recent examination (9.9%). |
| Khoo | 2016 | 2 | Random forest classifier | NR | 0.566 | 0.590 | NR | NR |
| Brandt | 2019 | 2 | Logistic | NR | 0.52 for | 0.54 for | The cancerous | The ipsilateral breast |
|  |  |  | regression |  | VPD, 0.53 for DV | VPD, 0.56 for DV | (ipsilateral) breast VPD decreased 0.26% and the contralateral breast VPD decreased 0.39% for a difference of 0.13%. For DV, the ipsilateral breast decreased 2.10 cm<sup>3</sup> and the contralateral breast decreased 2.74 cm<sup>3</sup> for a difference of 0.63 cm<sup>3</sup>. | VPD decreased 0.29% and the contralateral breast VPD decreased 0.28% for a difference of -0.02%. For DV, the ipsilateral breast decreased 1.82 cm<sup>3</sup> and the contralateral breast decreased 1.89 cm<sup>3</sup> for a difference of 0.05 cm<sup>3</sup>. |
| Tan | 2016 | 4 | Support vector machine based risk model | NR | 0.730 for prior 1, 0.710 for prior 2, 0.666 for prior 3 | NR | NR | NR |
| Vachon | 2007 | For cases, 5.0. For controls, 5.2. | Logistic regression and general estimating equations | NR | NR | 0.65 | Difference in percent density from earliest to latest mammogram: for the ipsilateral side, mean (SE) = -1.3% (7.5). For the contralateral side, mean (SE) = -1.5% (7.4). The difference in PD in the contralateral breast between cases and controls was 5.5% at 9 years before the cancer, 5.3%, at 5 years, and only 4.0% at 3 years. The corresponding values for the ipsilateral side were 4.9%, 4.9%, and 4.1%, respectively. | Difference in percent density from earliest to latest mammogram: for the ipsilateral side, mean (SE) = -1.2% (6.3). For the contralateral side, mean (SE) = -1.1% (6.5). The difference in PD in the contralateral breast between cases and controls was 5.5% at 9 years before the cancer, 5.3%, at 5 years, and only 4.0% at 3 years. The corresponding values for the ipsilateral side were 4.9%, 4.9%, and 4.1%, respectively. |

\*Kerlikowske 2015 uses change in density to predict future risk of breast cancer, whereas other studies report association between change in mammographic characteristics and risk of breast cancer

These studies show modest improvement in estimating 5- and 10-year risk with AUC increasing from 0.635 to 0.640 after adding change in density.^19^ Brandt shows similar modest change in AUC to discriminate cases from controls using volumetric percent density change in cancerous breast and normal breast from 0.52 to 0.54 though the time horizon appears to be the time between the two mammograms used for this study (median time 3 years).^14^ Tan on the other hand evaluated bilateral asymmetry of breast density between left and right breast as a marker of near term cancer risk.^29^

The statistical methods used to model change and assumptions including breast imaged (ipsilateral or contralateral to the cancer) and approach to comparing cases and controls for the other association studies are summarized in Table 4. Some studies used change in BIRADs category while others had continuous breast density generated from machine derived measures.

**Table 4.** Analytical models used for repeated measures of mammographic features that do not report AUC (sorted by number mammograms used)

| Author | Year | Avg number of mmgs used | Model of change (e.g., difference between two densities, mixed effects model, etc.) | Prediction horizon (5/10yr) | Risk other than AUC | Change for cases | Change for controls |
| --- | --- | --- | --- | --- | --- | --- | --- |
| Kerlikowske | 2007 | 2 | Multivariable logistic regression | NR | OR of breast cancer if BI-RADS breast density category increased from 1 to 2 (= 1.9) and 1 to 3 (= 3.4) | A total of 19.6% of all women had an increase in breast density category and 18.5% had a decrease. The majority of women had a BI-RADS breast density category of 2 or 3 on the first and last examination: 29.4% of women with breast cancer had BI-RADS scores of 2 on the first and last screens, and 24.3% women with breast cancer had scores of 3 on both screens. | A total of 19.6% of all women had an increase in breast density category and 18.5% had a decrease. The majority of women had a BI-RADS breast density category of 2 or 3 on the first and last examination: 30.0% of women without breast cancer had BI-RADS scores of 2 on the first and last screens, and 22.9% of women without breast cancer had scores of 3 on both screens. |
| Work | 2014 | 2 | Multi-variable conditional logistic regression and linear regression | NR | A >5% decrease in percent density was inversely associated with breast cancer (OR=0.56 for fully adjusted model), while a >5% increase in percent density was positively associated with breast cancer (OR=2.55); however, these associations were not statistically significant. | Percent density = +0.29% per year. Mean change = - 2.67% (range of - 34–16). | Percent density = -1.62% per year. Mean change = - 5.35% (range of -48–49). |
| Humphrey | 2016 | 2 | Paired t-tests, general estimating equations, and an exchangeable correlation matrix | NR | No statistically significant differences between cases and controls | PD change for affected breast = -0.4%, PD change for unaffected breast = -0.3% | PD change = -0.4% and -0.5% |
| Byrne | 2017 | 2 | Logistic regression | NR | Controlling for baseline mammographic density, a 1% change in mammographic density increased breast cancer risk 4%, but not statistically significantly, in women assigned placebos (OR = 1.04). The increase in breast cancer risk was not statistically significant (OR = 1.20) comparing the highest to the lowest quintile of mammographic density change. | For all women assigned placebo, mean mammographic density change = -0.05% with median of 0.0% | For all women assigned placebo, mean mammographic density change = -0.05% with median of 0.0% |
| Román | 2019 | 2 | Poisson regression | NR | Women whose density category increased from B to C or B to D had a RR of 1.55 and | Most frequently, women remained at density category B at earliest and latest examination | Most frequently, women remained at density category B at earliest and latest examination (40.8%). The proportion of women |
|  |  |  |  |  | 2.32, respectively. The RR for women whose density increased from C to D was 1.51. | (33.1%). The proportion of women that remained at BI-RADS density C or D was greater for women with breast cancer. 34.0% experienced a decrease and 12.5% experienced an increase in breast density category. | that remained at BI-RADS density A or B was significantly greater for women without breast cancer. 25.8% had a decrease, and 11.8% had an increase in breast density category. |
| Kim | 2021 | 2 | Poisson distribution | 5 year | For women with BI-RADS Category 4 during both screenings, the 5-year risk was 1.24%. | 23.0% of all women had a higher density category, and 22.2% had a lower density category in the second screening compared to the first screening. | 23.0% of all women had a higher density category, and 22.2% had a lower density category in the second screening compared to the first screening. |
| Kim | 2020 | NR | Flexible parametric proportional hazards models | NR | Multivariable-adjusted HRs for incident breast cancer comparing the regressed, developed, and persistent breast density groups with the “none” group were 1.81, 1.47, and 3.01, respectively. | NR | NR |
| Kang | 2021 | NR | Proportional hazards regression model | NR | In comparison to consistently nondense group, HR for consistently dense group = | For all women, 199 (5.5%) experienced a density decrease and 185 (5.1%) a density increase pre to | For all women, 199 (5.5%) experienced a density decrease and 185 (5.1%) a density increase pre to postmenopause. The other 89.4% of participants |
|  |  |  |  |  | 2.31 for fully adjusted model, HR for density decrease = 0.83, and HR for density increase = 1.04. In addition, compared to the participants with decreased breast density after menopause, participants with increased breast density had a four-fold greater risk of BC (HR = 4.27). | postmenopause. The other 89.4% of participants exhibited no changes in density; 641 (17.8%) had consistently dense breast tissue and 2,572 (71.5%) never exhibited dense breast. | exhibited no changes in density; 641 (17.8%) had consistently dense breast tissue and 2,572 (71.5%) never exhibited dense breast. |
| Salminen | 1998 | 2-5 | Multivariate analysis with the Cox proportional hazard model | NR | The age-adjusted relative risks (RR) of breast cancer among women with unfavourable parenchymal patterns of the breast at the first round were only marginally increased (RR varied from 1.5 to 1.3 among women with P2 to DY patterns). After taking into account the mammographic parenchymal pattern sequentially at the rounds preceding the diagnosis, the | At the first screening round, the prevalence of normal mammographic parenchymal pattern (N1) was 13% and the prevalence of DY pattern was about 4%. There was a drift in the mammographic parenchymal patterns from the unfavourable P2,DY types to the favourable N1,P1 types between the first and the last screening rounds. At the last screening round | At the first screening round, the prevalence of normal mammographic parenchymal pattern (N1) was 13% and the prevalence of DY pattern was about 4%. There was a drift in the mammographic parenchymal patterns from the unfavourable P2,DY types to the favourable N1,P1 types between the first and the last screening rounds. At the last screening round the prevalence of N1 patterns was 46% and that of DY patterns under 1%. |
|  |  |  |  |  | RRS varied from 2.6 to 4.7 and were statistically significant. There was only a small and not statistically significant increase in the risk of breast cancer among those women whose breast patterns changed either from favorable to unfavorable (RR = 1.3) or from unfavorable to favorable (RR = 1.2) compared with women whose patterns remained favorable. | the prevalence of N1 patterns was 46% and that of DY patterns under 1%. |  |
| Azam | 2020 | 2-5 | Cox proportional hazards regression | NR | Compared with women with a decreased MD over time, no statistically significant difference in BC risk was seen for women with either stable MD or increasing MD (hazard ratio = 1.01 and 0.98, respectively). Among premenopausal<br>women, there was a weak but statistically nonsignificant association between annual increase in MD greater than 10% and risk of BC (HR = 1.12) compared with premenopausal women with an annual decrease in MD greater than 10%.<br>Women ages 40-49 with an increase in annual MD greater than 10% had a statistically nonsignificant 30% higher risk compared to perimenopausal women with greater than 10% annual MD reduction. | NR | NR |
| Maskarinec | 2006 | 2.8 | Multilevel regression and linear regression | NR | The rate of change in percent density was not significantly related to case status (p = 0.11). | Unadjusted percent densities differed by ~20% between age 40 and 60. We estimated the age-related decline as 5.63% per 10 years. The nonlinear effect of 1.64% per 10 years in the full model<br>described the faster decline of densities over time earlier in life than later. The mean size of the total breast area was 25% larger at age 75 to 80 than at age 40 to 45, whereas the size of the dense areas decreased by 34% with age. | Unadjusted percent densities differed by ~20% between age 40 and 60. We estimated the age-related decline as 5.63% per 10 years. The nonlinear effect of 1.64% per 10 years in the full model described the faster decline of densities over time earlier in life than<br>later. The mean size of the total breast area was 25% larger at age 75 to 80 than at age 40 to 45, whereas the size of the dense areas decreased by 34% with age. |
| Sartor | 2020 | 3 | Wilcoxon signed-rank test | NR | We detected a statistically significant difference in breast density change over time ($p = 0.045$). | Density change = - 0.3% | Density change = +1.7% |
| van Gils | 1999 | 5 | Conditional logistic regression | 10 year | In women with 5–25% density initially, we observed a trend of decreasing risk with diminishing density: when women with <5% density throughout the whole period formed the reference category, the odds ratio (OR) for those who decreased from 5–25% to <5% density was 1.9 in contrast to the OR of 5.7 for those with persisting 5–25% density. In women who increased from 5–25% density to >25% density the OR was 6.9. | Majority of cases stayed at 5-25% density | Majority of controls stayed at 5-25% density |
| Lokate | 2013 | 5 | Linear mixed models and logistic regression | NR | For each percentage point decrease in mammographic density, odds ratio = 1.01. Those who increased in density by one or more categories seemed to have a slightly increased risk (statistically significant only for those increasing from the first to the second quartile of dense area, OR = 2.8). | The mean decline in percent density between the first and last available mammograms was 10.8% for both breast cancer cases and controls. The change in absolute dense area was not different for breast cancer cases and controls (mean = -15.2 cm<sup>2</sup>). The mean change in absolute nondense area was +12.3 cm<sup>2</sup>. 47% of all women showed a decline; 49% stayed in the same category; and 4% showed an increase in percent density over an average period of 10 years. Generally, women in whom breast density decreased with one or more categories had a slightly lower risk of breast cancer than did those who stayed in the same category, although not statistically significant. | The mean decline in percent density between the first and last available mammograms was 10.8% for both breast cancer cases and controls. The change in absolute dense area was not different for breast cancer cases and controls (mean = -15.2 cm<sup>2</sup>). The mean change in absolute nondense area was +7.3 cm<sup>2</sup>. 47% of all women showed a decline; 49% stayed in the same category; and 4% showed an increase in percent density over an average period of 10 years. Generally, women in whom breast density decreased with one or more categories had a slightly lower risk of breast cancer than did those who stayed in the same category, although not statistically significant. |
| Vachon | 2007 | For cases, 5.0. For controls, 5.2. | Logistic regression and general estimating equations | NR | For contralateral PD, the ORs range from 1.0043 (for change in PD of -10 or quartile 1) to 0.9972 (for change in PD of +6.5 or quartile 4). These results were similar for the ipsilateral side [0.9997 (for change in PD of -10 or quartile 1) to 1.0002 (for change in PD of +6.5 or quartile 4)]. The best-fitting model for both the ipsilateral and contralateral sides included no variables in which effects changed with follow-up (all P values testing for time-varying effects of mammographic density were 0.10 or higher). | Difference in percent density from earliest to latest mammogram: for the ipsilateral side, mean (SE) = -1.3% (7.5). For the contralateral side, mean (SE) = -1.5% (7.4). The difference in PD in the contralateral breast between cases and controls was 5.5% at 9 years before the cancer, 5.3%, at 5 years, and only 4.0% at 3 years. The corresponding values for the ipsilateral side were 4.9%, 4.9%, and 4.1%, respectively. | Difference in percent density from earliest to latest mammogram: for the ipsilateral side, mean (SE) = -1.2% (6.3). For the contralateral side, mean (SE) = -1.1% (6.5). The difference in PD in the contralateral breast between cases and controls was 5.5% at 9 years before the cancer, 5.3%, at 5 years, and only 4.0% at 3 years. The corresponding values for the ipsilateral side were 4.9%, 4.9%, and 4.1%, respectively. |
| Busana | 2016 | For cases, 5. For controls, | Linear mixed models and conditional logistic | NR | Women with a high PD at baseline, which remained high | NR | Cumulus = -1.17% a year, Image-J = -1.07% a year. The linear component of the yearly rate of change in |
|  |  | 7. | regression |  | over time, had the highest odds of developing BC relative to a woman with mean random intercept and mean slope (OR: 8.10 for Cumulus (left MLO) and 3.42 for the ImageJ-based method (left–right MLO mean). In contrast, women with the lowest PD at baseline, despite a slight increase in their PD over time, had the lowest odds of developing BC (OR: 0.07 for Cumulus (left MLO) and 0.23 for the ImageJ-based method (left–right MLO mean). |  | PD was more than twice as fast after the menopausal transition than prior to it for Cumulus (-1.10 % vs. -0.50 %, respectively). Similarly, the yearly rate coefficient in the ImageJ-based model was nearly twice as fast after the menopause (-1.16 % vs. 0.67 %, respectively). |

## Discussion

We identified 20 studies addressing change in mammographic breast density or other features and risk of breast cancer. Of these, 9 had only 2 images giving only modest ability to detect an association between change in density and risk of breast cancer. Only 6 studies report AUC for their analysis, and 5 of these use this measure to summarize discrimination of the cases from the controls. Only Kerlikowske uses change in density categories from BIRADs classification to predict 5- and 10-year risk. In the study, adding change in density to the prediction model gave a modest improvement in model performance. Overall, approaches to analysis of repeated mammogram images reflect the underlying approach to density (categorical or continuous) and this variation further limits interpretation of this body of evidence.

Focus of these studies is predominantly on mammographic breast density with limited study of change in texture features. Only 2 studies look at change in texture features.^21 29^ A recent meta-analysis of change in density and breast cancer risk used data from 4 cohort studies and reported a pooled HR for increase in breast density compared to women with non-dense breast tissue (HR = 1.61; 95% CI 1.33-1.92) for studies reporting hazard ratios and pooled OR those reporting odds ratios (OR = 1.98; 95% CI 1.31-3.0).^12^ In that meta-analysis, decrease in breast density was associated with reduced risk compared to women with stable breast density (HR = 0.78; 95% CI 0.71-0.87). Of note, a single study contributed multiple measures of change in density within this analysis without adjustment for use of a common reference group.

Interval between images used for change is quite variable (range from as short as 1 year to median of 4.1 years). The majority of studies evaluate change in category of density; for example, BIRADs not a continuous measure of density. With only 2 images used, change in category is limited and the shorter time interval between images reduces power to differentiate trajectories of mammographic features over time. We might ask, given the low rate of decline in mammographic density with age as described,^34 35^ is the interval used in these studies sufficient to detect meaningful change? To address this gap in the literature, future studies should use repeated measures methods incorporating more mammographic images over longer time periods.

There is a steady decline in mammographic breast density through midlife to menopause and beyond.^34 35^ This slow decrease over time makes a discrete change in category harder to capture and will be limited compared to use of continuous mammographic density measures that are now becoming more broadly available. Future studies using the continuous density measures may better capture change and the risk associated with these changes.

While breast cancer rarely develops simultaneously in both breasts, current models still utilize average mammographic density and/or other features between the two breasts in conducting the risk prediction. Although mammographic density from the two breasts appears to be highly correlated at the baseline, deviation between the two breasts may be better captured over time using repeated mammography. Based on this review of the literature, we conclude that longitudinal bivariate analysis^36^ of mammograms has never been used in breast cancer epidemiology.

We note limited use of change measures for improving risk prediction. Prediction to next routine mammogram may reflect available evidence but it is not helpful for current risk reduction recommendations for high-risk women (lifestyle changes, chemoprevention, or surgery), each typically with a longer term 5- or 10-year time horizon.^5 37^

We live in a precision medicine society. For high-income countries, this requires translation of advances in biotechnology to focus treatment and prevention according to level of risk, and further balance risks and benefits of treatment or prevention.^38^ Cancer prevention is often conceptualized as strategies that interrupt cancer pathways and maximize the short- and long-term benefits of prevention intervention.^39-41^ To implement precision prevention, we need refined strategies for 5- or 10-year risk classification^38^ that can be applied in real time in the clinical setting, such as in the context of screening mammography which remains a standard for early detection of BC.^4 42^ Focus, therefore, should be placed on better use of repeated mammographic measures of breast features to stratify risk and identify both the high-risk groups and also the low-risk groups^43^ to tailor screening and prevention strategies.^44^

## Limitations

There are several limitations with the current review. Heterogeneity of the data did not allow for a meta-analysis. Additionally, systematic reviews are always subject to possible publication bias if all relevant studies have not been published. We used several strategies to reduce the risk of this including using a thorough search strategy designed by a medical librarian with expertise in searching for systematic reviews, and searching clinicaltrials.gov for any ongoing studies.

## Conclusion

Despite current limitations in the literature, the more widespread use of digital mammography and availability of digital images repeated over time offers growing opportunities to improve risk classification and risk prediction for women.

## Data Availability

All data produced in the present study are available upon reasonable request to the authors

## Complete Search Strategies

### Search strategies designed and executed by Angela Hardi, MLIS Embase.com

=3,919 results on 9/9/2020 (Limited to English; editorials, letters, and notes excluded from results)

**Updated search** (date limited to 2020-present): 602 on 10/14/2021 (‘breast density’/exp OR ((breast NEAR/3 densit*):ti,ab,kw OR (mammary NEAR/3 densit*):ti,ab,kw OR (mammographic NEAR/3 densit*):ti,ab,kw)) AND (‘mammography’/de OR mammograph*:ti,ab,kw OR mammogram*:ti,ab,kw OR mastrography:ti,ab,kw OR ‘digital breast tomosynthesis’:ti,ab,kw OR ‘x-ray breast tomosynthesis’:ti,ab,kw) NOT (‘editorial’/it OR ‘letter’/it OR ‘note’/it) AND [english]/lim

### Ovid Medline All

= 2694 results on 9/9/2020 (Limited to English; editorials, comments, and letters excluded) **Updated search** (date limited to 2020-present): 440 results on 10/14/2021 (Breast Density/ OR (breast adj3 densit*).ti,ab. OR (mammary adj3 densit*).ti,ab. OR (mammographic adj3 densit*).ti,ab.) AND (Mammography/ OR mammograph*.ti,ab. OR mammogram*.ti,ab. OR mastrography.ti,ab. OR “digital breast tomosynthesis”.ti,ab. OR “x-ray breast tomosynthesis”.ti,ab.) NOT (comment.pt. OR editorial.pt. OR letter.pt.)

### CINAHL Plus

=978 results on 9/9/2020; (Limited to English and these publication types: Clinical Trial, Corrected Article, Journal Article, Meta Analysis, Meta Synthesis, Practice Guidelines, Proceedings, Protocol, Randomized Controlled Trial, Research, Review, Systematic Review)

**Updated search** (dated limited to 2020-present): 135 results on 10/14/2021 ((MH “Breast Tissue Density”) OR AB(breast N3 densit*) OR TI(breast N3 densit*) OR AB(mammary N3 densit*) TI(mammary N3 densit*) OR AB(mammographic N3 densit*) OR TI(mammographic N3 densit*)) AND ((MH “Mammography”) OR AB(mammograph*) OR TI(mammograph*) OR AB(mammogram*) OR TI(mammogram*) OR AB(mastrography) OR TI(mastrography) OR AB(“digital breast tomosynthesis”) OR TI(“digital breast tomosynthesis”) OR AB(“x-ray breast tomosynthesis”) OR TI(“x-ray breast tomosynthesis”))

### Scopus

=3,162 results on 9/9/2020 (Limited to English; editorials, notes, letters, and book chapters excluded from results)

**Updated search** (date limited to 2020-present): 423 results on 10/14/2021 TITLE-ABS ((breast W/3 densit*) OR (mammary W/3 densit*) OR (mammographic W/3 densit*)) AND TITLE-ABS (mammograph* OR mammogram* OR mastrography OR “digital breast tomosynthesis” OR “x-ray breast tomosynthesis”) AND (EXCLUDE (DOCTYPE, “no”) OR EXCLUDE (DOCTYPE, “ch”) OR EXCLUDE (DOCTYPE, “le”) OR EXCLUDE (DOCTYPE, “ed”)) AND (LIMIT-TO (LANGUAGE, “English”))

### Cochrane Library

=358 results on 9/9/2020 (1 Cochrane Protocol and 357 results from CENTRAL Trials)

**Updated Search** (date limited 2020-present in CENTRAL Trials) = 32 results on 10/14/2021

ID Search

#1 MeSH descriptor: [Breast Density] explode all trees

#2 ((breast NEAR/3 densit*) OR (mammary NEAR/3 densit*) OR (mammary NEAR/3 densit*) OR (mammographic NEAR/3 densit*)):ti,ab,kw

#3 #1 OR #2

#4 MeSH descriptor: [Mammography] explode all trees

#5 (mammograph* OR mammogram* OR mastrography OR “digital breast tomosynthesis” OR “xray breast tomosynthesis”):ti,ab,kw

#6 #4 OR #5

#7 #3 AND #6

### ClinicalTrials.gov

= 11 results (searched the “Other terms” field) on 9/9/2020

**Updated Search** = 12 results on 10/14/2021 (1 new result, added to the Excel library)

(“breast density” OR “mammary density”) AND (mammograph* OR mammogram* OR mastrography OR “digital breast tomosynthesis” OR “x-ray breast tomosynthesis”)

